# Pulmonary Fibrosis after COVID-19 is Associated with Severity of Illness and Blood Leukocyte Telomere Length

**DOI:** 10.1101/2021.03.17.21253834

**Authors:** Claire F. McGroder, David Zhang, Mohammad A. Choudhury, Mary M. Salvatore, Belinda M. D’Souza, Eric A. Hoffman, Ying Wei, Matthew R. Baldwin, Christine Kim Garcia

**Author notes:** Corresponding author: Christine Kim Garcia, MD, PhD, Professor of Medicine, Columbia University Irving Medical Center; New York, NY; 10032,. Drs. Baldwin and Garcia served as co-senior authors.

## Abstract

The risk factors for development of fibrotic interstitial lung abnormalities (ILA) after severe COVID-19 are incompletely described and the extent to which CT findings correlate with symptoms and physical function after hospitalization remain unclear. At 4 months after hospitalization, fibrotic ILA was more common in those who underwent mechanical ventilation (72%) than in those who did not (20%). We demonstrate that severity of initial illness, duration of mechanical ventilation, lactate dehydrogenase on admission, and leukocyte telomere length are independent risk factors for fibrotic ILA. These fibrotic changes correlate with lung function, cough and measures of frailty, but not with dyspnea.

## INTRODUCTION

Reports of hospitalized COVID-19 survivors show that there are persistent symptoms, radiographic abnormalities, and physiologic impairments months after the initial illness^1,2^. Persistent chest imaging abnormalities and histopathologic findings of lung fibrosis were also found in a majority of survivors of the SARS-CoV-1 2003 outbreak^3,4^, suggesting that the SARS viruses may lead to a worse fibroproliferative response than other pneumonias.

Cohort studies of COVID-19 survivors report that severity of the initial illness is associated with a greater risk of persistent CT abnormalities^1,2,5,6^, especially for patients requiring supplemental oxygen or mechanical ventilation, but independent clinical, biomarker, and genomic risk factors have not been identified. Also, the extent to which CT findings correlate with symptoms and physical function remain unclear. To address knowledge gaps, we conducted a prospective cohort study of survivors hospitalized with severe COVID-19, half of whom were mechanically ventilated, with four month follow-up. We sought to characterize associations of pulmonary radiographic and physiologic sequela of severe COVID-19, and to identify independent risk factors for the development of post-COVID fibrosis.

## METHODS

### Additional details are included in the Supplementary Materials

We conducted a single-center prospective cohort study of adults hospitalized between March 1, 2020 and May 15, 2020 who required supplemental oxygen. At four months after hospitalization, participants underwent a non-contrast high resolution chest CT scan (HRCT), pulmonary function testing, measurement of six-minute walk distance (6MWD) and assessments of frailty. Radiographic interstitial lung abnormalities (ILAs) were categorized as suggested by established criteria^7^, quantitated using a severity scoring system developed by ARDSnet and used in ARDS patients survivors^8,9^, and binned into two groups (non-fibrotic and fibrotic). Fibrotic ILAs included those with reticulations, traction bronchiectasis and honeycombing. Genomic DNA was isolated from blood collected at follow-up.

We calculated Spearman’s rank correlation coefficients between continuous data. We created separate generalized additive logistic models (GAMs) to test adjusted associations between the risk of fibrotic ILA with independent continuous variables identified in univariable analysis. Due to the moderate cohort size and rate of fibrotic ILA, we used generalized covariate balanced propensity scores (CBPS) to adjust for potential confounders. We estimated adjusted odds ratios using logistic regression models if there was no evidence of non-linearity in the GAMs.

## RESULTS

We enrolled 76 patients meeting eligibility criteria (**Figure S1**); demographic and clinical features are shown in **Table S1**. All participants required supplemental oxygen during hospitalization, and 32 (42%) required mechanical ventilation.

A median 4.4 (IQR: 4.0-4.8) months after hospitalization, the most common radiographic abnormality was ground glass opacities (43%), followed by reticulations (39%) and traction bronchiectasis (28%) (**Figure 1, Table S2**). Fibrotic ILA’s were more common in those who were mechanically ventilated compared to those who were not (72% vs. 20%, p=0.001) (**Table S3**). In unadjusted analyses, those with fibrotic ILA were significantly more likely to be male, have shorter telomeres, higher admission SOFA scores, higher lactate dehydrogenase levels, and have received steroids, or anti-IL6 receptor blockade (**Table S1**). Qualitative measures of non-fibrotic and fibrotic ILA were closely associated with quantitative scores (**Figure S2**).

**Figure 1.**
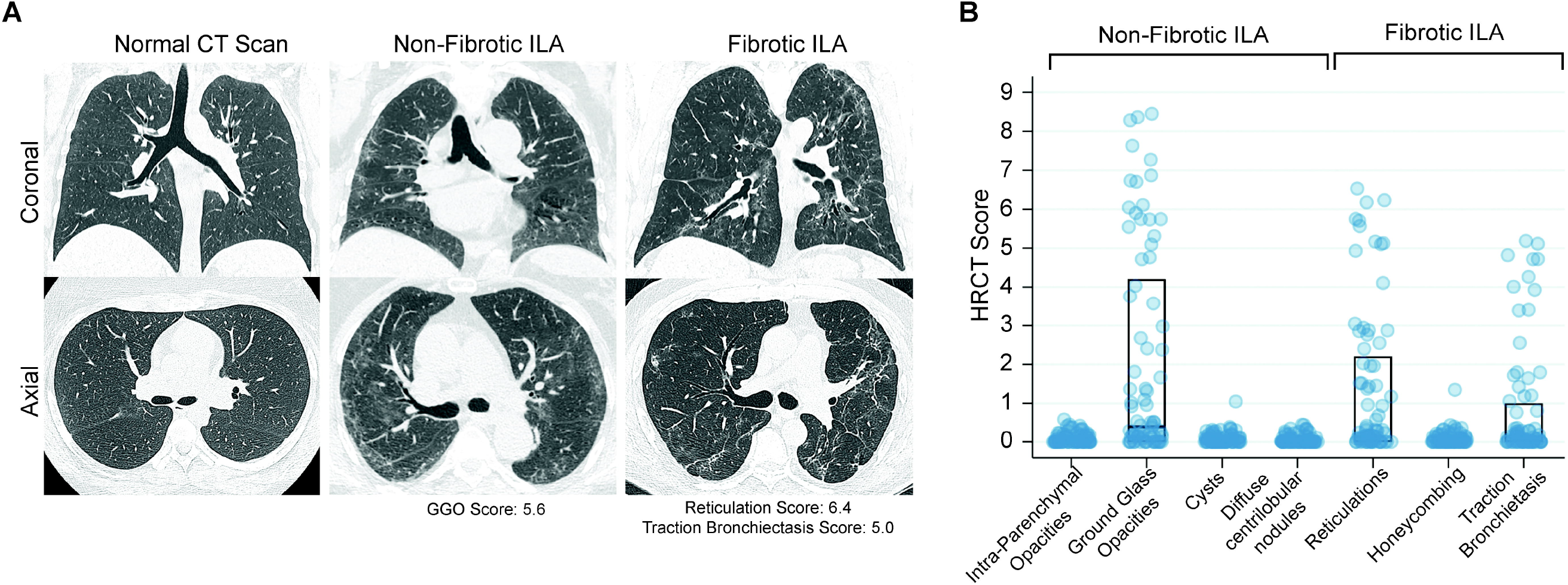
High resolution Computed Tomography (CT) scans of the chest from COVID-19 Survivors. **(A)** Representative CT chest scans demonstrating no abnormalities (left), non-fibrotic Interstitial Lung Abnormalities (ILA) (middle), and fibrotic ILA (right). The upper panels show a coronal section and the lower panels show an axial image at the level just below the carina. The non-fibrotic ILA scan had a Ground Glass Opacities (GGO) score of 5.6 (84^th^ percentile in the group). The fibrotic ILA scan had a reticulation score of 6.4 (98^th^ percentile), a traction bronchiectasis score of 5.0 (95^th^ percentile), and no honeycombing. (**B)** Chest high-resolution computed tomography (HRCT) scores for ILA categories of abnormalities observed in the study cohort. The middle line of box plot represents the median score; bottom and top lines represent the 25^th^ and 75^th^ percentile, respectively. Where no lines are seen the 25^th^, 50^th^, and 75^th^ percentile scores were all 0. Severity of ILA was graded using a scoring system developed by ARDSnet^8,9^. The possible range of scores was 0-20 from all categories of abnormalities, except traction bronchiectasis, which had a possible range of 0-5.

Participants had an array of functional deficits (**Table S4**). Overall, 40 (53%) had a reduced diffusion capacity, 78% had a decreased 6MWD, 18% remained >10% below baseline weight, and 53% had weak grip strength.

Ground glass, reticulations, and traction bronchiectasis scores correlated more strongly with reduction in diffusion capacity (*ρ* −0.34, −0.64, and −0.49, respectively, all p<0.01) than forced vital capacity (**Table 1**). Ground glass correlated with frailty, while reticulation and traction bronchiectasis correlated with cough. Dyspnea correlated with frailty score, grip strength, and 6MWD, but not radiographic patterns (**Table 1**).

**Table 1.** Spearman Correlation Coefficients of Radiographic and Dyspnea scores with Pulmonary Function, and 6-Minute Walk Distance, Frailty and Symptoms.

**A.**
| CT Pattern | DLCO (% predicted) |  |  | FVC (% predicted) |  |  | 6MWD (m) |  |  |
| --- | --- | --- | --- | --- | --- | --- | --- | --- | --- |
|  | R<sup>2</sup> | r | p | R<sup>2</sup> | r | p | R<sup>2</sup> | r | p |
| Ground Glass Opacities | 0.12 | -0.34 | **0.003\*** | 0.06 | -0.25 | **0.03\*** | 0.00 | -0.02 | 0.92 |
| Reticulations | 0.41 | -0.64 | **<0.001\*** | 0.04 | -0.21 | 0.07 | 0.00 | -0.02 | 0.80 |
| Traction Bronchiectasis | 0.24 | -0.49 | **<0.001\*** | 0.05 | -0.23 | **0.04\*** | 0.00 | -0.05 | 0.69 |
| CT Pattern | Frailty Phenotype Score |  |  | Cough Scale |  |  | UCSD SOBQ |  |  |
|  | R<sup>2</sup> | r | p | R<sup>2</sup> | r | p | R<sup>2</sup> | r | p |
| Ground Glass Opacities | 0.21 | 0.46 | **<0.001\*** | 0.00 | 0.07 | 0.56 | 0.02 | 0.14 | 0.23 |
| Reticulations | 0.05 | 0.23 | **0.04\*** | 0.07 | 0.26 | **0.02\*** | 0.00 | 0.05 | 0.66 |
| Traction Bronchiectasis | 0.03 | 0.16 | 0.17 | 0.06 | 0.25 | **0.03\*** | 0.00 | 0.07 | 0.57 |

**B.**
|  | DLCO (% predicted) |  |  | FVC (% predicted) |  |  | 6MWD (m) |  |  |
| --- | --- | --- | --- | --- | --- | --- | --- | --- | --- |
|  | R<sup>2</sup> | r | p | R<sup>2</sup> | r | p | R<sup>2</sup> | r | p |
| UCSD SOBQ | 0.02 | -0.14 | 0.24 | 0.06 | -0.25 | **0.04\*** | 0.06 | -0.25 | **0.03\*** |
|  | Frailty Phenotype Score |  |  | Grip Strength (kg) |  |  | Gait Speed (m/s) |  |  |
|  | R<sup>2</sup> | r | p | R<sup>2</sup> | r | p | R<sup>2</sup> | r | p |
| UCSD SOBQ | 0.22 | 0.47 | **<0.001\*** | 0.14 | -0.37 | **0.001\*** | 0.05 | -0.21 | 0.06 |

Abbreviations: DLCO, diffusion capacity for carbon monoxide; FVC, forced vital capacity; 6MWD, six-minute walk distance; CT, computed tomography. UCSD: University of California San Diego Shortness of Breath Questionnaire
\* Significant after controlling for false discovery using the Benjamini-Hochberg method at an FDR of 0.10.

Fully-adjusted GAM’s showed that both admission SOFA score and percent-predicted telomere length were linearly associated with the predicted risk of fibrotic ILA (**Figure 2**). Duration of mechanical ventilation varied linearly with the predicted risk of fibrotic ILA through 20 days and plateaued with more prolonged mechanical ventilation. LDH levels also plateaued at higher levels. In fully-adjusted logistic regression models, every 1 point increase in SOFA score, 50-point increase in LDH, and 1 ventilator-day was associated with 1.49 (95% CI 1.17 − 1.89), 1.24 (95% CI 1.08 − 1.43), and 1.07 (95% CI 1.03 - 1.12), higher odds of fibrotic ILA, respectively. Each 10% decrease in age-adjusted LTL was associated with a 1.35 higher odds of fibrotic ILA (95% CI 1.06- 1.72). Sensitivity analyses are shown in **Figure S3** and **S4**.

**Figure 2.**
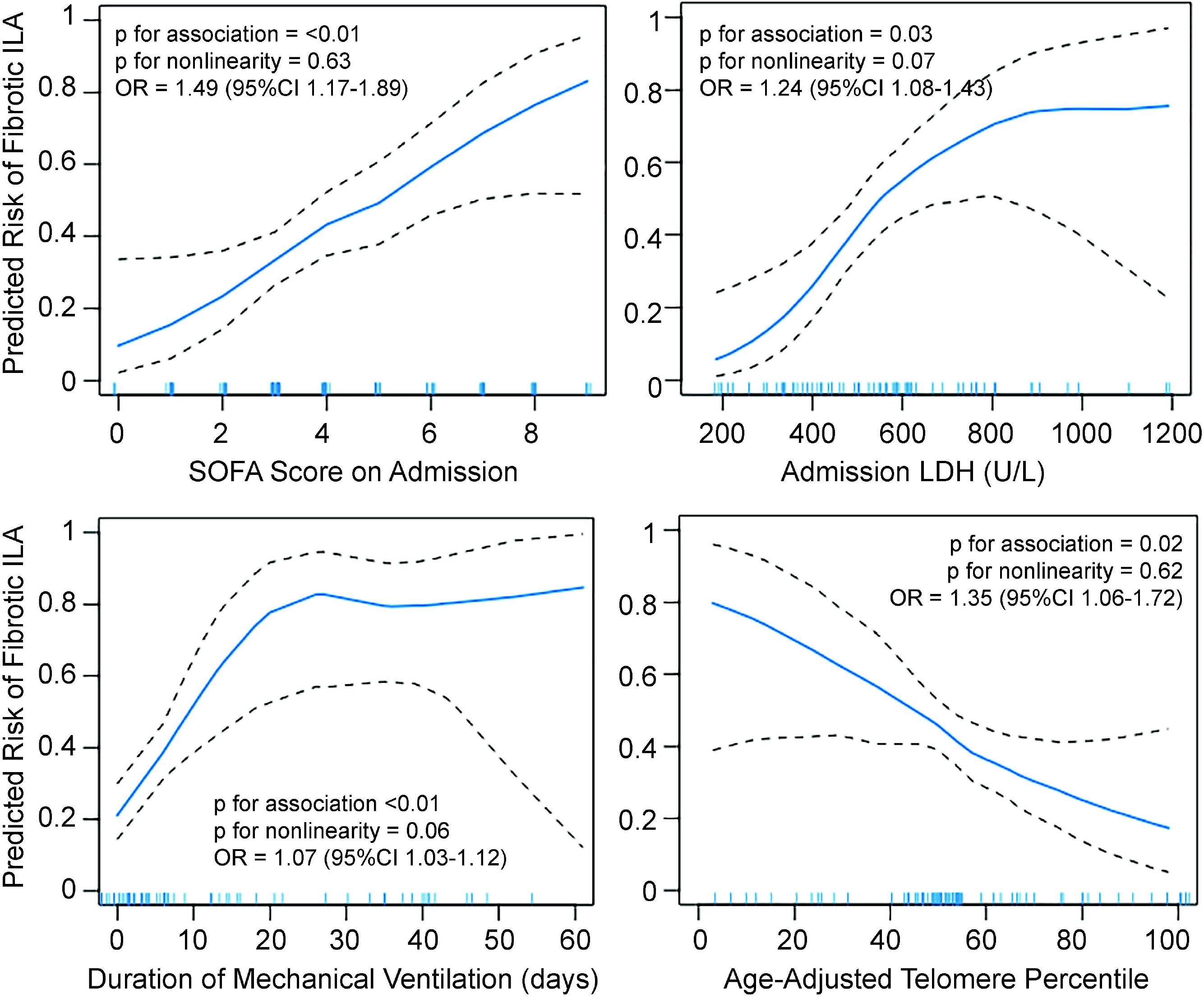
Continuous associations of fibrotic interstitial lung abnormalities (ILA) with admission Sequential Organ Failure Assessment (SOFA) score (top left), LDH levels (top right), duration of mechanical ventilation in days (bottom left), and age-adjusted leukocyte telomere length percentile (bottom right) using generalized additive models with LOESS smothers. Blue line represents predicted values. Black dashed lines are 95% confidence intervals. Hash marks along the x-axis indicate individual study participants. Since there was no evidence for non-linearity in the generalized additive models, we estimated adjusted odds ratios using logistic regression models. All models are adjusted for age, sex, race/ethnicity, days since infection, body mass index, pack-years of smoking, treatment with steroids while hospitalized, and the other independent variables of interest. Odds ratio for SOFA score is per point increase, for LDH is per 50 point increase, for mechanical ventilation is per day increase, and for telomere length is per 10% decrease in percent-predicted value.

## DISCUSSION

Pulmonary fibrosis is a feared complication of respiratory infections. We found that 20% of non-mechanically ventilated and 72% of mechanically ventilated COVID-19 survivors had fibrotic ILAs four months after hospitalization. The presence of these radiographic abnormalities correlates with decrements in lung function, cough, and frailty. Greater initial severity of illness, longer duration of mechanical ventilation, and shorter blood leukocyte telomere length are each independent risk factors for the development of fibrotic ILA.

Fibrosis was measured in this study both subjectively, in a manner congruent with other ARDS^8^, population-based^10^ and COVID-19^1,2,6^ studies, as well as objectively using texture analysis^11^. We include reticulations as a manifestation of a fibrotic ILA to facilitate comparison to prior ARDS and SARS CoV1 literature^4,9^ and acknowledge that, although a prevalent radiographic finding after infection, reticulations may either resolve or progress over time. However, the presence of the persistent pulmonary function degradations and radiographic findings at four months are concerning for potential long-term damage.

This is the first study to identify age-adjusted LTL as an independent risk factor for post-COVID pulmonary fibrosis. Short blood leukocyte telomere lengths have been shown to be a risk factor for the development of different subtypes of fibrotic interstitial lung disease (ILD), including IPF^12,13^. Here, we also find that longer telomere lengths appear to be protective, with less fibrotic ILA following severe infection. Thus, this genomic biomarker may measure the balance of pro- and anti-fibrotic susceptibilities.

Limitations of this study include its small size, the lack of replication cohort, and the possibility that COVID-19 may itself affect telomere length. Our primary outcome, ILA, may precede COVID infection. Patients were hospitalized prior to FDA-approved therapies, yet half received steroids.

Still, this study reveals significant respiratory symptoms and morbidity associated with severe COVID-19. Dyspnea, as noted by most survivors^14^, correlates more strongly with muscle strength and frailty than ILAs, suggesting persistent multisystemic dysfunction. Additional prospective studies are needed to characterize temporal changes of post-COVID-19 fibrotic abnormalities, and clinical trials are needed to investigate therapeutic options to promote its resolution.

## Data Availability

The authors confirm that the data supporting the findings of this study are available within the article and its supplementary materials. Raw data were generated at Columbia University Medical Center. Derived data supporting the findings of this study are available from the corresponding author [CG] on request.

## ACKNOWLEDGEMENTS

The authors wish to thank the patients for their participation, Deanna D. Rivas and Mason W. Amelotte for excellent technical assistance, and the Safety Monitoring Board (SMB) Chair Joao De Andrade, MD, and SMB members Anil Vachani, MD and Anna Rozenshtein, MD. This work was supported by grants from the NIH (R01HL103676, R01HL093096 to CKG; T32HL105323 to CM; UL1TR001873 to MRB) and the Department of Defense (PR202907 to CKG and MRB). The study sponsors had no role in data analysis and interpretation, manuscript writing, or dissemination of results.

