## Supplement for "Pulmonary Fibrosis after COVID-19 is Associated with Severity of Illness and Blood Leukocyte Telomere Length"

**Supplemental Methods.**

**Table S1. Demographic and clinical features of COVID-19 survivors.**

**Table S2. Prevalence of HRCT abnormalities.**

**Table S3. Clinical Features of COVID-Survivors who underwent mechanical ventilation.**

**Table S4. Prevalence of CT abnormalities, lung function and physical impairment, and respiratory symptoms.**

**Table S5. Spearman correlation between SOFA score, ventilator days, and age-adjusted percent telomere length and covariables before and after covariate-balancing propensity score.**

**Figure S1. Study Flow Diagram**

**Figure S2. Continuous Association of percent Ground Glass Opacity (GGO) and Ground Glass Reticulation (GGR) via AMFM lung texture analysis and Any ILA and Fibrotic ILA.**

**Figure S3. Sensitivity analysis of the continuous association of fibrotic interstitial lung abnormalities (ILA) with SOFA score, lactate dehydrogenase, days of mechanical ventilation, and age-adjusted leukocyte telomere length (LTL) percentile using generalized additive models with LOESS smothers.**

**Figure S4. Sensitivity analysis of the continuous association of fibrotic interstitial lung abnormalities (ILA) as defined by Fleishner Society position paper with SOFA score, lactate dehydrogenase, days of mechanical ventilation, and age-adjusted leukocyte telomere length (LTL) percentile using generalized additive models with LOESS smothers.**

**Supplemental Methods.**

*Study Design*

We conducted a single-center prospective cohort study of adults age 21 years and older hospitalized between March 1, 2020 and May 15, 2020 with a positive SARS-CoV-2 RT-PCR nasopharyngeal swab and who required supplemental oxygen. The Columbia University Irving Medical Center (CUIMC) Institutional Review Board study protocol number is AAAR1916. Participants signed a written informed consent.

*Recruitment and Enrollment*

We enrolled participants by calling via telephone consecutive patients meeting eligibility criteria based on their admission date, with sampling weighted to include approximately 50% survivors of mechanical ventilation. We sought to enroll 70-100 participants based upon the capacity of the research team to recruit and assess participants during the 4-month follow-up study period of July and August 2020. Participants ambulated independently prior to hospitalization, did not live in a skilled-care facility prior to hospitalization, required supplemental oxygen therapy during their hospital stay, and were discharged to acute rehabilitation, subacute rehabilitation, or home. All participants were living at home prior to enrollment. We excluded those with pre-existing interstitial lung disease and those who had undergone lung transplantation.

*Electronic Medical Record Measurements*

We obtained clinical data from the New York Presbyterian-CUIMC clinical data warehouse, which contains electronic data for inpatient and outpatient visits[1]. Patient data included demographics, diagnoses, procedures, medications, laboratory tests, vital signs and ventilator flowsheet data, and other clinical variables. Past medical history diagnoses were retrieved from the hospital admission notes and by using groups of ICD-10 diagnosis codes according to the Clinical Classifications Software by the Healthcare Cost and Utilization Project[2]. We calculated the Sequential Organ Failure Assessment (SOFA) score during the first 24 hour of admission[3].

*Chest Computed Tomography*

Non-contrast high resolution chest CT scans (HRCT) were performed at maximal inspiration using either GE VCT 64 or GE Revolution CT750 HD instrument. Two chest radiologists (MS and BD) evaluated the scans for radiographic abnormalities, using interstitial lung abnormality (ILA) categories recommended by a recent Fleishner position paper[4] (ground glass opacities, non-emphysematous cysts, reticulations, honeycombing, and traction bronchiectasis) in addition to centrilobular nodules and intra-parenchymal opacities, which have been described in COVID-19 patients[5,6]. The radiographic abnormalities were scored using a semi-quantitative scoring system used by ARDSnet investigators[7]. Those with traction bronchiectasis, honeycombing, or reticulations were categorized as having a fibrotic ILA. All others were categorized as having a non-fibrotic ILA. Radiographic abnormalities were assessed at 5 levels: the aortic arch, 1cm above the diaphragm, and three levels equally spaced between the aortic arch and diaphragm. Each level was divided into four lung quadrants, and scored within each quadrant as: 0=no involvement; 1= <5% involvement; 2= 5–25% involvement; 3=26–49% involvement; 4=50–75% involvement; and 5=greater than 75% involvement. The sum of the quadrant scores at each level was averaged across the five levels to determine the final score (range 0-20). For traction bronchiectasis, an airway abnormality that is difficult to quantify, each level was scored as either 0 or 1 for its absence or presence, respectively, for a maximum score of 5.

In addition to the radiologists' subjective scoring of ILAs, the University of Iowa imaging lab (Iowa City, IA, USA) used the adaptive multiple features method (AMFM) to quantify HRCT scans for various lung features, including: ground glass opacity, ground glass-reticular, honeycombing, emphysema, or normal lung[8].

*Clinical Measurements*

Pulmonary function tests were performed on one of two pulmonary function machines (NDD EasyOne Pro, Andover MA; Vyaire Medical VMAX Encore, Mettawa IL). Pulmonary function testing and six-minute walk distance (6MWD) were assessed according to established guidelines[9,10]. Cough was assessed using a 100mm visual analogue scale[11]. Dyspnea was assessed using the UCSD shortness of breath questionnaire[12].

*Frailty Measurements*

We measured the five Fried Frailty domains: gait-speed, grip-strength, weight loss, low activity, and exhaustion. We measured grip-strength, gait-speed, and exhaustion, and using the traditional Cardiovascular Health Study (CHS) methodology[13]. Weight loss was calculated as the difference between their hospitalization admission weight and the measured weight during the follow up visit. A decrease >10 lbs. met the Fried Frailty domain requirement. We assessed the physical activity domain on the basis of report of activities performed at four-moth follow-up using the Duke Activity Status Index (DASI)[14] instead of the the Minnesota Leisure Time Physical Activity Questionnaire[15], the original CHS measure of physical activity, as it improves the construct and predictive validity of frailty assessments in ARF survivors[16]. We used previously validated DASI score cutoffs for low activity in older ARF survivors (men $\leq$12.5; women $\leq$10)[16].

Each frailty domain is assigned 1 point if present and 0 points if absent based on the established cutoffs (range, 0-5). Consistent with CHS methodology, we defined the frailty phenotype as being frail in $\geq$3 of the five domains.

*Genetic and Genomic Measurements*

Blood was collected from each participant at the follow-up visit. DNA was isolated from blood leukocytes using the Gentra Puregene Blood kit (Qiagen, Valencia CA). Leukocyte telomere length (LTL) was measured using a quantitative PCR assay and the RotoGene real-type PCR system (Qiagen)[17]. The LTL was expressed as a logarithm-transformed ratio of telomere to single-copy gene [ln(T/S)] and this value was compared to LTL from normal control subjects (n=201 unrelated multiethnic individuals from Dallas, TX, ranging in age from 19 to 89 years) to estimate an age-adjusted LTL percentile.

*Missing Data*

The Glasgow Coma Score, a component of the SOFA score, was missing for nearly all non-ICU patients, and we thus imputed a score of 15 for these participants based on previous literature[18]. Inflammatory markers ESR, CRP, LDH, Ferritin and D-dimer, had 6-12% missingness in a pattern that was not completely random. We used the MICE function in R(v3.5.1) to perform multiple imputation using predictive mean matching for missing values. There were six participants that could not perform the 6-minute walk test because they were non-ambulatory and one whose baseline heart rate was too high to safely perform the test. For the six participants who were non-ambulatory, we imputed 0 and ran a complete case analysis excluding the last participant. Three participants were unable to produce acceptable or reproducible DLCO measure. Given low degree of missingness, we ran a complete case analyses using only participants with complete PFT data.

*Statistical Analyses*

We examined unadjusted associations of clinical and biomarker characteristics with no ILA, non-fibrotic ILA, or fibrotic ILA using analysis of variance (ANOVA), Kruskal Wallis, or chi-squared tests. We calculated correlation coefficients between continuous data using

Spearman's method. We examined adjusted associations of fibrotic ILA with the hypothesized and biologically plausible independent variables that were associated with fibrotic ILA in unadjusted analyses. Due to the moderate cohort size and rate of fibrotic ILA, we used generalized covariate balanced propensity scores (CBPS) to adjust for potential confounders so as to avoid model overparameterization, as has been done in prior studies[19]. We generated propensity scores predicting each of the independent variables of interest using the CBPS function in R(v3.5.1)[20]. To do so, we calculate CBPS for each independent variable by regressing it on a set of potential confounders age, sex, race/ethnicity, days since infection, body mass index, pack-years of smoking, treatment with steroids while hospitalized, and the two other independent variables of interest. We created separate generalized additive logistic models (GAMs) for each independent continuous variable. The resulting CBPS was then included in the GAM model as an additional covariate. In the GAMs, we did not assume linear associations between continuous independent variables and risk of fibrotic ILA, and instead estimated associations by a nonparametric locally weighted smoothing spline. For lab values like LDH, we removed outliers beyond 2 standard deviations for the analysis. For independent variables without evidence of non-linear associations with fibrotic ILA in the GAMs, we estimated adjusted odds ratios using logistic regression models. Since LTL may increase severity of illness[21], and since LTL may be affected by critical illness[22], we conducted sensitivity analyses: one estimating associations of LTL, admission SOFA score, LDH, and duration of mechanical ventilation with fibrotic ILA without adjusting for the other three independent variables. The second sensitivity analysis was performed with a more conservative definition of fibrotic ILA that comprises only traction bronchiectasis and honeycombing[4]. Covariates were found to be balanced in all propensity scores (see **Table S5**)[23,24]. Analysis were performed using Stata/IC v16 (StataCorp) and R, v3.5.1.

**Table S1. Demographic and clinical features of COVID-19 survivors.**

| | Total | None | Non-Fibrotic ILA | Fibrotic ILA | p-value* |
|---|---|---|---|---|---|
| Number | 76 | 31 (41%) | 13 (17%) | 32 (42%) | |
| DEMOGRAPHICS | | | | | |
| Age, mean (SD) | 54.0 (13.7) | 51.8 (13.7) | 60.6 (10.8) | 53.4 (14.2) | 0.14 |
| Male (%) | 45 (61%) | 16 (52%) | 4 (31%) | 25 (78%) | **0.007** |
| Hispanic ethnicity | 43 (57%) | 14 (45%) | 10 (77%) | 19 (59%) | 0.14 |
| Race | | | | | 0.17 |
| White | 30 (39%) | 13 (42%) | 4 (31%) | 13 (41%) | |
| Black | 22 (30%) | 13 (42%) | 4 (31%) | 5 (16%) | |
| Asian | 1 (1%) | 0 | 0 | 1 (4%) | |
| Other | 23 (30%) | 5 (16%) | 5 (38%) | 13 (41%) | |
| Body Mass Index (kg/m$^2$), mean (SD) | 32.2 (6.9) | 34.2 (7.8) | 32.7 (4.6) | 30.1 (6.4) | 0.07 |
| GENOMIC FACTORS | | | | | |
| Leukocyte telomere length, percentile (IQR) | 52 (49-68) | 52 (50-81) | 52 (52-82) | 49.5 (40-52) | **0.01** |
| LTL <10th percentile, N | 3 (4%) | 0 | 0 | 3 (9%) | 0.11 |
| COMORBIDITIES | | | | | |
| Hypertension | 41 (54%) | 20 (65%) | 5 (38%) | 16 (50%) | 0.24 |
| Diabetes | 25 (33%) | 12 (41%) | 6 (46%) | 7 (22%) | 0.20 |
| COPD | 4 (5%) | 1 (3%) | 0 | 3 (9%) | 0.36 |
| Asthma | 18 (27%) | 8 (26%) | 4 (31%) | 6 (19%) | 0.65 |
| Heart Disease | 2 (3%) | 1 (3%) | 1 (8%) | 0 | 0.17 |
| Chronic Kidney Disease | 7 (9%) | 5 (16%) | 0 | 2 (6%) | 0.29 |
| Smoking Status | | | | | |
| Ever | 31 (41%) | 16 (52%) | 4 (31%) | 11 (34%) | 0.27 |
| Active | 2 (3%) | 1 (3%) | 0 | 1 (2%) | 0.81 |
| Pack Years (IQR) | 0 (0-10) | 1 (0-15) | 0 (0-1) | 0 (0-9) | 0.34 |
| CLINICAL FACTORS | | | | | |
| Admission SOFA Score, mean (SD) | 4.1 (2.4) | 2.9 (1.7) | 4.5 (3.4) | 5.3 (2.4) | **0.001** |
| Received Steroids | 39 (51%) | 10 (32%) | 5 (38%) | 24 (75%) | **0.002** |
| Received Anti IL-6R blocker** | 17 (22%) | 3 (10%) | 1 (8%) | 13 (41%) | **0.005** |
| Venous thromboembolism (by imaging) | 12 (16%) | 4 (13%) | 1 (8%) | 7 (22%) | 0.42 |
| Received Therapeutic Anticoagulation | 25 (33%) | 6 (19%) | 3 (23%) | 16 (50%) | **0.03** |
| Maximum Oxygen Requirement | | | | | **<0.001** |
| Nasal Cannula | 23 (30%) | 18 (58%) | 4 (31%) | 1 (3%) | |
| Non-Rebreather | 17 (22%) | 6 (19%) | 5 (38%) | 6 (19%) | |
| NIPPV or HFNC | 4 (5%) | 2 (7%) | 0 | 2 (6%) | |
| Mechanical Ventilation | 31 (41%) | 5 (16%) | 4 (31%) | 22 (69%) | |
| MV plus ECMO | 1 (1%) | 0 | 0 | 1 (3%) | |
| Ventilator Days (IQR) | 30.5 (12-42) | 7 (6-9) | 32.5 (20-41) | 34 (14-42) | **<0.001** |
| Hospital days (IQR) | 18 (7-35) | 4 (4-12) | 16 (9-26) | 35 (24-54) | **<0.001** |
| Discharge Disposition | | | | | **0.001** |
| Home | 54 (71%) | 29 (94%) | 10 (77%) | 15 (47%) | |
| Acute Rehabilitation | 19 (25%) | 1 (3%) | 2 (15%) | 16 (50%) | |
| Subacute Rehabilitation | 3 (4%) | 1 (3%) | 1 (8%) | 1 (3%) | |
| Home-dwelling at 4 months | 76 (100%) | 31 (100%) | 13 (100%) | 32(100%) | 1.00 |
| OUTCOMES | | | | | |
| Oxygen use at 4 months | 4 (5%) | 1 (3%) | 2 (15%) | 1 (3%) | 0.25 |
| FVC, % predicted, mean (SD) | 83.6 (19.7) | 87.6 (21.9) | 83.2 (13.6) | 79.8 (19.2) | 0.19 |
| DL(CO), % predicted, mean (SD) | 74.9 (42.1) | 90.5 (24.5) | 74.2 (16.7) | 60.5 (16.6) | **<0.001** |
| 6MWD, m, mean (SD) | 364 (98) | 355 (105) | 370 (72) | 370 (103) | 0.82 |
| 6MWD, % predicted, mean (SD) | 68 (18) | 67 (19) | 80 (18) | 65 (16) | 0.06 |
| Change in weight,kg, † median (IQR) | -0.2 (-5.1 - 2.3) | 1.2 (-0.9 - 5.2) | -2.1 (-4.6 - 1.7) | -3.5 (-11.6 - 0.3) | **<0.001** |

Data are n (%) unless otherwise specified.

Abbreviations: ILA, Interstitial Lung Abnormalities; COPD, Chronic Obstructive Pulmonary Disease; LDH, lactate dehydrogenase; IL-6R, interleukin 6 receptor; NIPPV, non-invasive positive pressure ventilation; HFNC, high-flow nasal cannula, MV, mechanical ventilation; ECMO, extracorporeal membrane oxygenation; FVC, forced vital capacity; DL(CO), diffusion capacity for carbon monoxide; 6MWD, 6 minute walk distance

* Associated examined using Chi-square, ANOVA or Kruskall-Wallis where appropriate

**Either tocilizumab or sarulimab

† Change in weight calculated as weight (kg) measured at 4 month visit minus weight documented at hospital admission.

**Table S2. Prevalence of HRCT Abnormalities.**

| | Total (n=76) | Non-Fibrotic ILA (n=13) | Fibrotic ILA (n=32) |
|---|---|---|---|
| Any Abnormality | 45 (59%) | | |
| Non-Fibrotic Abnormalities*: | | | |
| Intraparenchymal opacities | 1 (1%) | 0 | 1 (3%) |
| Ground glass opacities | 33 (43%) | 13 (100%) | 20 (62%) |
| Nonemphysematous cysts | 1 (1%) | 0 | 1 (3%) |
| Fibrotic Abnormalities*: | | | |
| Reticulations | 30 (39%) | 0 | 30 (94%) |
| Honeycombing | 1 (1%) | 0 | 1 (3%) |
| Traction Bronchiectasis | 21 (28%) | 0 | 21 (66%) |

HRCT: high resolution computed tomography; ILA: interstitial lung abnormalities

* patterns are not mutually exclusive and participants in the fibrotic ILA group can have non-fibrotic CT patterns as well as fibrotic patterns.

**Table S3. Clinical features of COVID-19 survivors who underwent mechanical ventilation.**

| | Total<br>N = 32 | No ILA<br>N = 5 | Non-Fibrotic ILA<br>N = 4 | Fibrotic ILA<br>N = 23 | p-value |
|---|---|---|---|---|---|
| **Ventilator support, Days** | 30.5 (12-42) | 7 (6-9) | 32.5 (20-41) | 34 (14-42) | **<0.001** |
| **$PaO_2$:$FiO_2$*, mean (SD)** | 166 (78) | 166 (57) | 165 (127) | 166 (76) | 0.99 |
| **Sequential Organ Failure Assessment (SOFA)** | 5.7 (2.3) | 3.4 (0.55) | 7.0 (0.82) | 5.9 (2.4) | **0.03** |
| **Positive End-Expiratory Pressure (cmH2O)*** | 15 (12-16) | 12 (12-14) | 14 (10-18) | 15 (14-16) | 0.33 |
| **Tidal Volume (per cc/kg ideal body weight)*** | 6.4 (5.9-7.3) | 6.2 (6.1-7.0) | 7.6 (5.6-8.1) | 6.4 (5.7-6.8) | 0.22 |
| **Received Steroids (%)** | 20 (62%) | 2 (40%) | 1 (25%) | 17 (74%) | 0.09 |
| **Prone Positioning (%)** | 9 (28%) | 0 | 0 | 9 (39%) | 0.09 |
| **Paralysis (%)** | 14 (44%) | 0 | 1 (25%) | 13 (57%) | **0.05** |

Abbreviations: ILA, interstitial lung abnormalities; PaO2, partial pressure of arterial oxygen; FiO2, Fraction of inspired oxygen

*These represent the first measured data following intubation

**Table S4. Prevalence of CT abnormalities, lung function and physical impairment, and respiratory symptoms.**

| Abnormality | N (%) |
|---|---|
| Any ILA | 45 (59%) |
| Fibrotic ILA | 32 (42%) |
| Reduced FVC* | 27 (36%) |
| Reduced FEV1/FVC* | 4 (5%) |
| Reduced DLCO* | 40 (53%) |
| Short 6MWD** | 59 (78%) |
| Weight Loss >10% baseline | 14 (18%) |
| Weight Loss >10 lbs† | 21 (28%) |
| Slow Gait Speed† | 18 (24%) |
| Weak Grip† | 40 (53%) |
| Decreased Activity† | 15 (20%) |
| Exhaustion† | 15 (20%) |
| UCSD SOBQ ≥ 10 | 52 (68%) |
| Cough Score ≥ 20‡ | 11 (14%) |

Abbreviations: ILA, interstitial lung abnormality; FVC, forced vital capacity; FEV1, forced expiratory volume in one second; DLCO, diffusion capac.y for carbon monoxide; 6MWD, six-minute walk distance; UCSD SOBQ, University of California San Diego Shortness of Breath Questionnaire.
*A reduced FVC or FEV1/FVC was defined as being below the lower limit of normal (LLN) for each subject.
** A short 6MWD was defined as being <80 percent of their predicted values.
† Fried frailty domain criteria (see Supplemental methods).
‡Cough visual analog score

**Table S5. Spearman correlation between SOFA score, ventilator days, and age-adjusted percent telomere length and covariables before and after covariate-balancing propensity score.**

| SOFA score, hospital admission | Pre-CBPS weighting | Post-CBPS weighting |
|---|---|---|
| Age | 0.027 | 0.140 |
| Sex | 0.316 | 0.080 |
| Time from COVID+ nasal swab to CT scan, in days | 0.220 | 0.166 |
| Black/African American | -0.187 | -0.027 |
| Hispanic | 0.249 | 0.037 |
| Other | -0.010 | -0.017 |
| BMI | -0.178 | -0.004 |
| Smoking, pack-years | 0.046 | 0.067 |
| Steroid therapy | 0.363 | 0.190 |
| Telomere length | -0.333 | -0.122 |
| Ventilator days | 0.531 | 0.166 |
| **Ventilator Days** | **Pre-CBPS weighting** | **Post-CBPS weighting** |
| Age | -0.189 | -0.166 |
| Sex | 0.326 | 0.159 |
| Time from COVID+ nasal swab to CT scan, in days | 0.293 | 0.128 |
| Black/African American | -0.218 | -0.064 |
| Hispanic | 0.235 | 0.128 |
| Other | 0.068 | -0.044 |
| BMI | -0.173 | -0.046 |
| Smoking, pack-years | -0.117 | -0.040 |
| Steroid therapy | 0.125 | 0.175 |
| Telomere length | -0.117 | -0.101 |
| SOFA score, day of admission | 0.532 | 0.273 |
| **Telomere Length (TL)** | **Pre-CBPS weighting** | **Post-CBPS weighting** |
| Age | -0.074 | -0.015 |
| Sex | -0.046 | -0.045 |
| Time from COVID+ nasal swab to CT scan, in days | -0.12 | -0.075 |
| Black/African American | -0.179 | 0.053 |
| Hispanic | -0.173 | -0.039 |
| Other | 0.088 | -0.001 |
| BMI | 0.106 | 0.061 |
| Smoking, pack-years | -0.117 | -0.006 |
| Steroid therapy | -0.142 | -0.035 |
| SOFA, day of admission | -0.333 | -0.108 |
| Ventilator days | -0.117 | -0.054 |

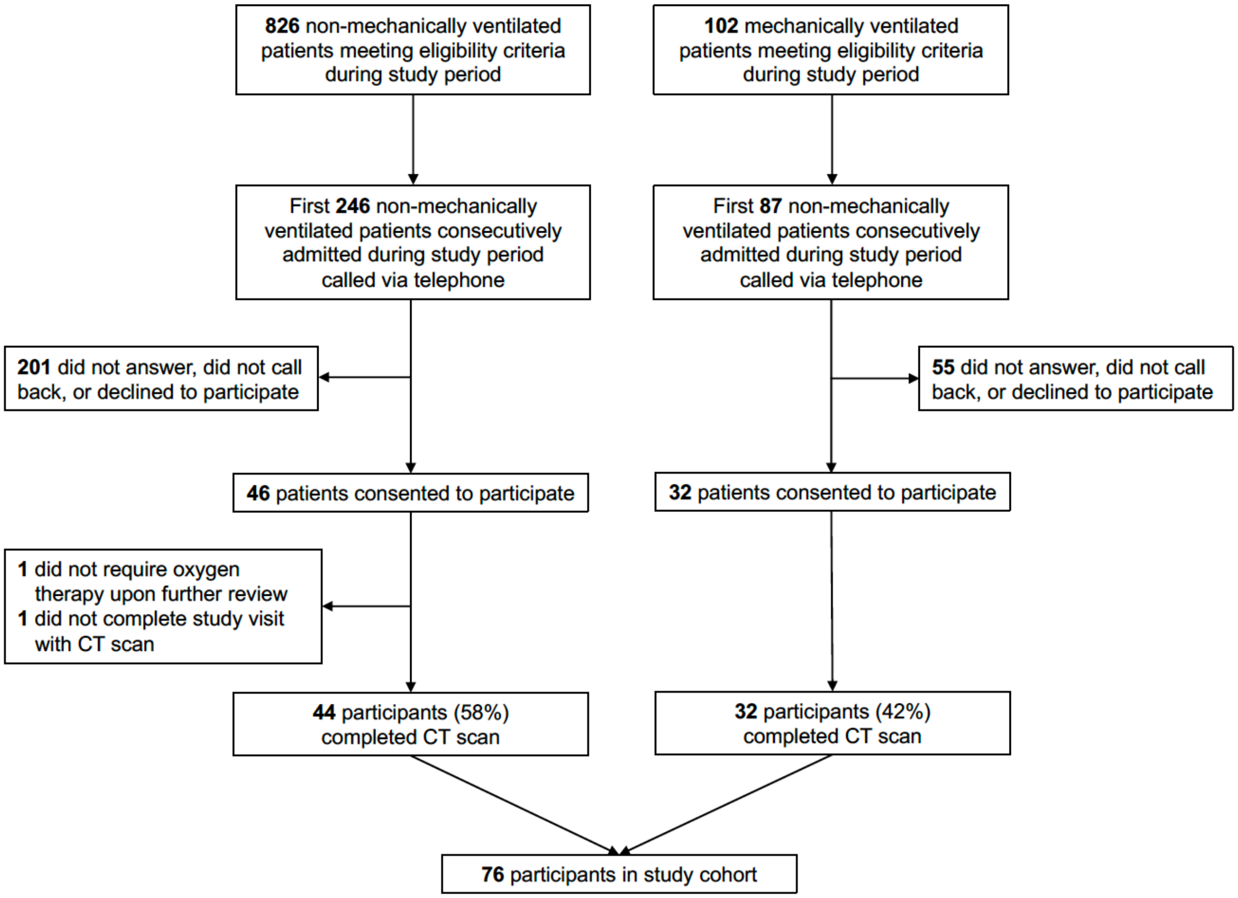


**Figure S1. Study Flow Diagram**

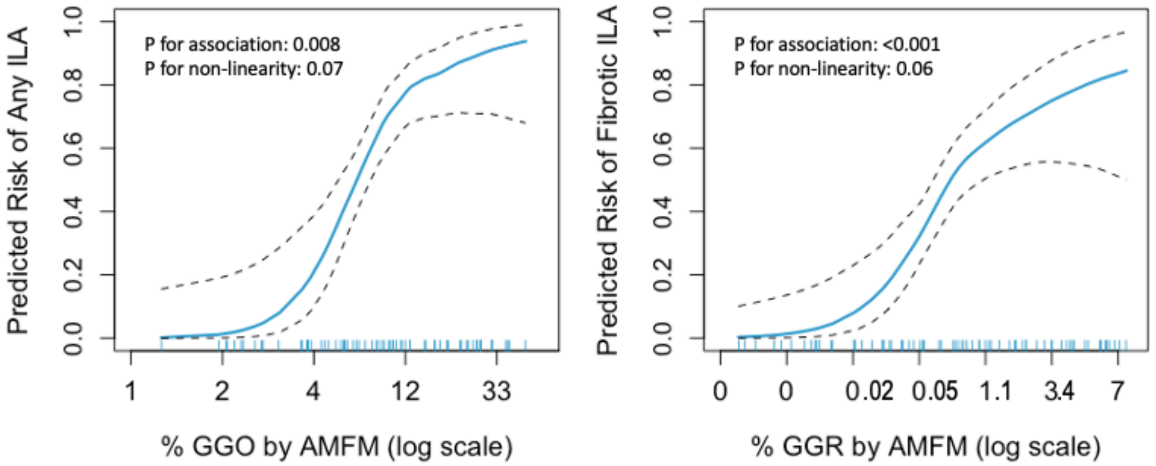


**Figure S2. Continuous Association of the natural log of the percent of Ground Glass Opacity (GGO) pattern as measured by Adaptive Multiple Features Method (AMFM) and any ILA (left) and percent of Ground Glass Reticulation (GGR) pattern via AMFM and fibrotic ILA (right) using generalized additive models with LOESS Smoothers. Each model is adjusted for age, sex, race/ethnicity and BMI. Both models are adjusted for age, sex, race/ethnicity and BMI.**

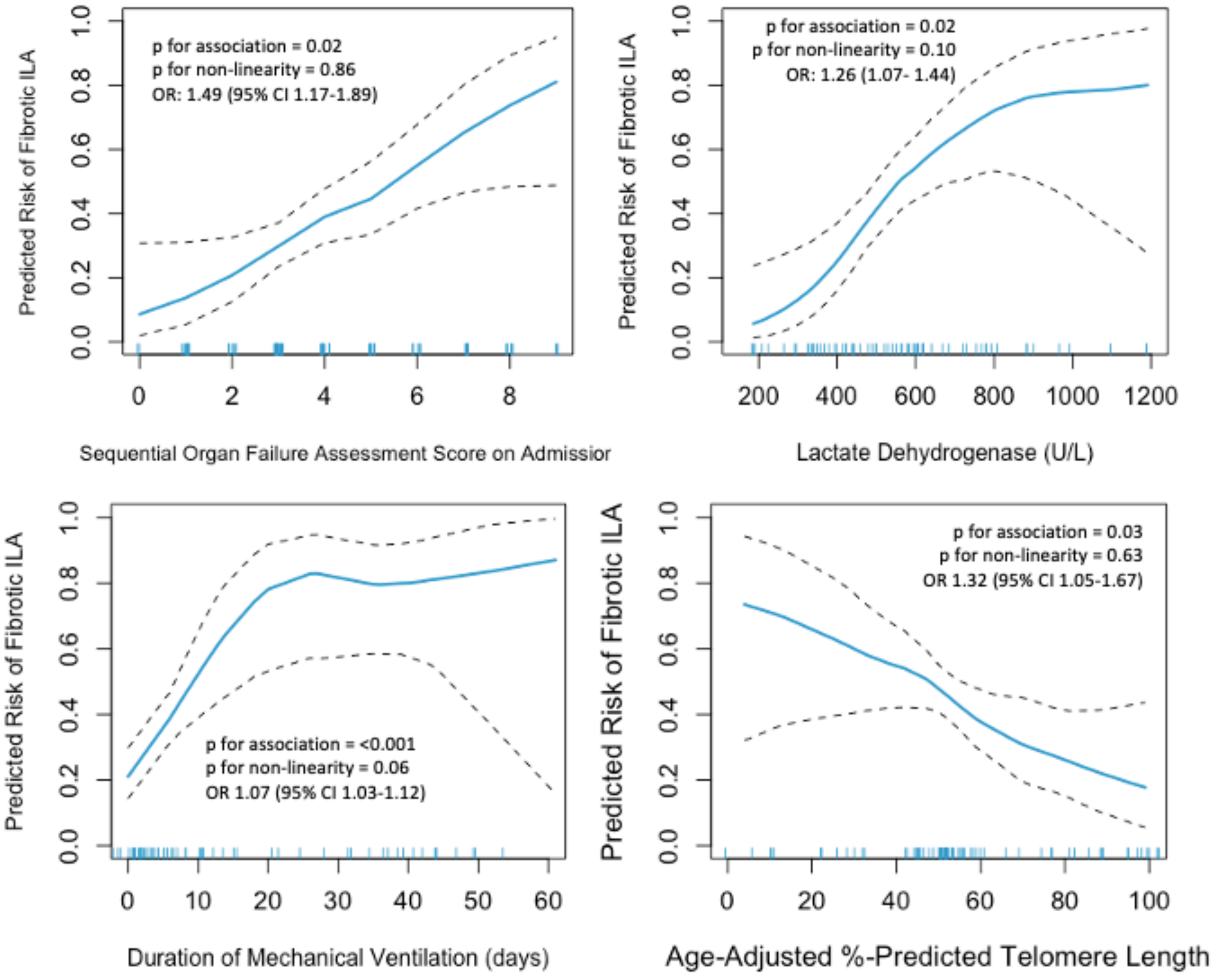


**Figure S3. Sensitivity of the Continuous Association of Fibrotic Interstitial Lung Abnormalities (ILA) with SOFA score (top left), Lactate Dehydrogenase (top right), Ventilator Days (bottom left) and Age-Adjusted Leukocyte Telomere Length Percentile (bottom right) using Generalized Additive Models with LOESS Smoothers. Each model is adjusted for age, sex, race/ethnicity, days since infection, body mass index, pack-years of smoking and treatment with steroids while hospitalized, but not the other three independent variables.**

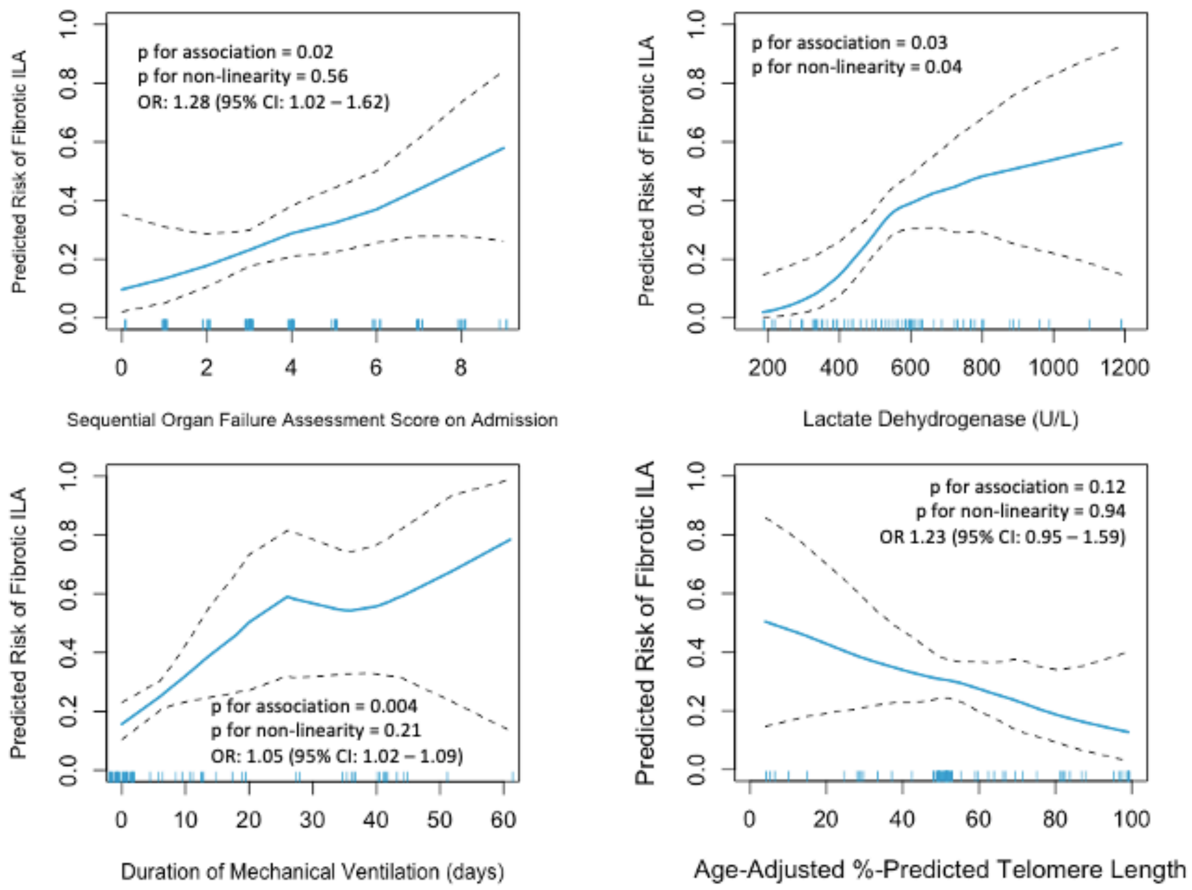


**Figure S4. Sensitivity of the continuous association of fibrotic Interstitial Lung Abnormalities (ILA) as defined by the presence of traction bronchiectasis or honeycombing (without reference to subpleural localization) as suggested by the ILA Fleishner Society position paper[4] with SOFA score (top left), Lactate Dehydrogenase (top right), Ventilator Days (bottom left) and Age-Adjusted Leukocyte Telomere Length Percentile (bottom right) using Generalized Additive Models with LOESS Smoothers. Each model is adjusted for age, sex, race/ethnicity, days since infection, body mass index, pack-years of smoking and treatment with steroids while hospitalized and the other three independent variables.**
